# A Continuous Bayesian Model for the Stimulation COVID-19 Epidemic Dynamics

**DOI:** 10.1101/2021.06.20.21259220

**Authors:** Zhaobin Xu, Hongmei Zhang, Yonggang Niu

## Abstract

It is of great theoretical and application value to accurately forecast the spreading dynamics of COVID-19 epidemic. We first proposed and established a Bayesian model to predict the epidemic spreading behavior. In this model, the infection probability matrix is estimated according to the individual contact frequency in certain population group. This infection probability matrix is highly correlated with population geographic distribution, population age structure and so on. This model can effectively avoid the prediction malfunction by using the traditional ordinary differential equation methods such as *SIR* (susceptible, infectious and recovered) model and so on. Meanwhile, it would forecast the epidemic distribution and predict the epidemic hot spots geographically at different time. According to the results revealed by Bayesian model, the effect of population geographical distribution should be considered in the prediction of epidemic situation, and there is no simple derivation relationship between the threshold of group immunity and the virus reproduction number *R*_0_. If we further consider the virus mutation effect and the antibody attenuation effect, with a large global population spatial distribution, it will be difficult for us to eliminate Covid-19 in a short time even with vaccination endeavor. Covid-19 may exist in human society for a long time, and the epidemic caused by re-infection is characterized by a wild-geometric && low-probability distribution with no epidemic hotspots.

## Introduction

As of June 1, 2021, the COVID-19 epidemic has caused 170 million infections and more than 3.6 million deaths worldwide, becoming the largest public health crisis facing the world after World War II. It is of great academic and practical value to predict the trend of COVID-19 epidemic situation. At this cutting edge, it is necessary to answer several questions urgently: whether the infection in Covid-19 can be completely eliminated by adopting group immunization, and what is the relationship between the threshold of group immunization and virus reproduction constant *R*_0_^[1,2]^.

However, the majority of our mathematical models if not all, are not be able to predict the epidemic trend well, although most of them have a good fitting result. A crucial reason behind this drawback, from our point of view, is the ignorance of the population geographic distribution. SIR model, which originated from epidemiological research, is a classic model to simulate infectious disease dynamics. It still occupies a central position in epidemiology, and the core lies in a set of ordinary differential equations. SIR model describes the relationship among three population groups under epidemic: susceptible, infectious and recovered. Without considering the spatial distribution characteristics of population, it is difficult to accurately estimate the development of epidemic situation by using the traditional SIR model. The initial epidemic development predicted by *SIR* model is often too dramatic, close to exponential shape, and the daily infections will gradually reach its peak with the consumption of susceptible population before a diminishing phase follows eventually ^[3-6]^. However, actual situation is beyond the expectations of any model. In order to better simulate infectious diseases, especially infections of Covid-19, we put forward a Bayesian model of virus infection for the first time. This model can take the information of population contact into account, so it can simulate the spreading dynamics more accurately. Not only that, this approach can integrate many features into the model, such as virus mutation factor, population age distribution, public prevention and control measures, etc., to further generate a more reliable prediction. This model can predict the epidemic development in real cases, and provide lots of valuable information for the development of the epidemic and the epidemiological tracking of infection cases.

## Methods

### 1.1 The establishment of individual contact matrix within a population

We have established a continuous Bayesian model of infection occurrence. Basic principles of this idea are briefly described below:

It is assumed that there are *N* individuals in a population, and there are different contact probabilities among those individuals. The infection probability is positively correlated with contact probability. For the simplified model, the relation constant is 1 which means the infection probability is equal to contact probability. The contact probabilities with themselves are zero. In this way, an *N*\**N* matrix is established, which has the following characteristics:

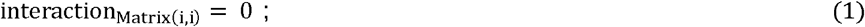

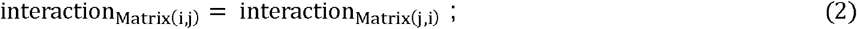

The accurate contact matrix can be obtained through tracking the individual contact probability in a real population group. For example, the position of each person’s mobile phone can be recorded to obtain the population contact matrix within certain time phase. The contact matrix is temporal and dynamic which means it will change though time. However, it is difficult for us to obtain such real data at present. Therefore, our model determines the contact frequency according to the relative distance between individuals. Therefore, our contact matrix is a theoretical and fixed contact matrix.

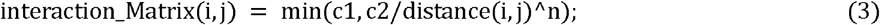

We assume that the contact probability between two different individuals is equal to the constant *c2* divided by the *n*th power of the distance between them with a upper bound equal to *c1*. We can preliminarily determine *c1, c2* and *N* according to the initial reproduction constant *R*_0_of virus.

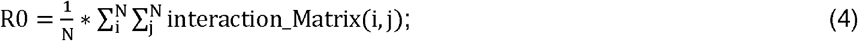

We further establish the Infection_Matrix with *N*\**M* elements, where *N* represents the population of the studied group and *M* represents the total number of generations.

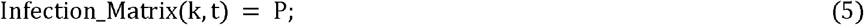

Equation(5) represents that the infection probability of individual *k* in the *t*-th infection generation is *P*.

### 1.2 A simplified Bayesian model

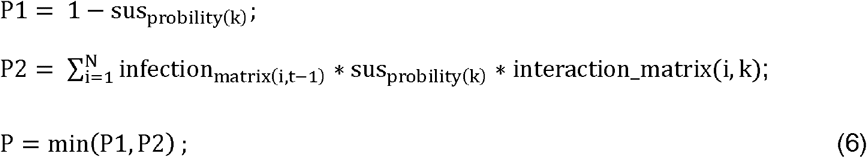

In which infection_matrix(i,t−1)_ represents the probability of infection of individual *i* in the previous generation, and sus_probility(k)_ represents the susceptibility constant of individual *k* in the *t*-th infection generation, which is between 0 and 1; interaction_matrix(i,k)represents the contact probability between individual *i* and individual *k*.

### 1.3 A Bayesian model considering complex factors

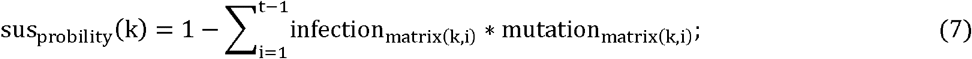

In which infection_matrix(k,i)_ represents the attenuation effect caused by virus mutation and antibody attenuation, and represents the attenuation effect of individual *k* in the *i*-th generation. If we think that there is no antibody attenuation and virus mutation effect, then infection_matrix(k,i)_ = 1.

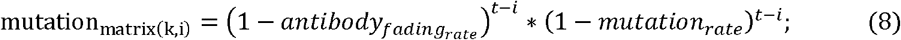

In which *t* represents the current virus generation or the current time, and 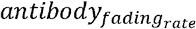 represents the attenuation constant of antibody with time (number of infected generations), because it represents the attenuation constant of a single generation, so it is a small number. Similarly, *mutation*_*rate*_ represents the variation constant of virus with time (number of infected generations), because it represents the variation constant during a single generation, so it is also a small number. Although the values of these two constants are small, the iteration effect of several generations will also cause a significant decrease in mutation_matrix(k,i)_. We need to make a rough estimate of these two parameters. Assuming that the average infection cycle in Covid-19 is 7 days, according to literature and news reports ^[7]^, the vaccine protection caused by Indian mutant *B*.*1*.*617*.*2* is about 1-88% = 0.12, and the Indian mutant occurs around the 50th infection cycle; The decline of vaccine protection caused by British mutant *B*.*1*.*1*.*7* is about 1-93% =0.07, and the occurrence time of British mutant is about the 30th infection cycle, so it is preliminarily inferred that mutation _ rate = 0.002; According to the statistical data of re-infection after infection, it shows that for people under 65 years old, the average protection rate of preventing the second infection after infection within 50 infection cycles is 80%. We speculate that the protection rate after 50 infection cycles is much lower than 80%, and it is calculated by 70% ^[8]^. Based on the mutation constants of viruses, we can preliminarily infer the antibody attenuation constant 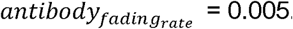.

#### Relationship between age and infection probability

A more accurate mathematical model should also take the influence of population’s immune variation into consideration. Our model mainly considers the influence of age-related immunity vibration on infection risk. According to the statistical results of infection distribution at different ages, the relationship between infection probability and age is further derived as equation(9).

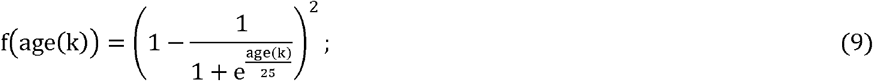

age(k) indicates the age of the *k*-th individual in the population.

#### Dose effects on infection

According to our research, the occurrence of infection is related to the initial number of virus invasions. Therefore, for people with low infection probability, the probability of becoming an infectious individual is less than the probability of producing antibodies, that is, the infected person does not necessarily have symptoms, or even positive reaction in nucleic acid test. Therefore, we added a correction function f(x) to express the relationship between the infection rate and the development of an individual into an infectious individual.

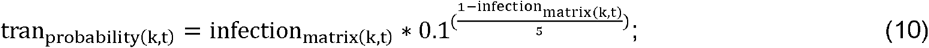

Therefore the final Bayesian model becomes:

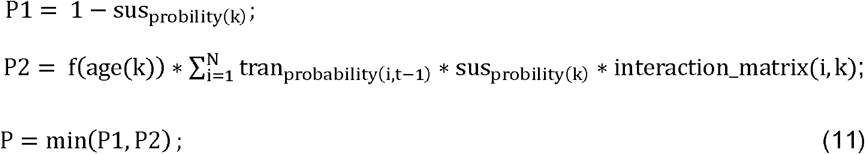

The virus reproduction coefficient *R*_0_ becomes:

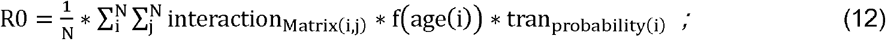

## 2 Results

### 2.1 An illustration of simple Bayesian model

The detailed description of the Bayesian model is explained in methods part. For the sake of better illustration, here we use a simple but concrete case to explain our model. Firstly, we study a simple Bayesian model without considering complicated factors. This model has the following assumptions

1. The individual immunity to certain infectious diseases is homogeneous, and there is no individual variation;
2. There is neither virus mutation nor the antibody attenuation effect with time;
3. All infections will have the same transmission potential, that is, if an individual is infected, it will produce antibodies, and at the same time it is contagious;
4. Individuals will recover after an infection cycle without death, that is, the overall population size will not change.

Taking this model as an example, we simply listed three individuals A, B, and C to study their infection possibility in the first few virus transmission cycles, assuming that the contact matrix between them is as follows:

According to this contact matrix, the initial virus reproduction coefficient *R*_0_ = 1/3 * (0+0.8+0.5+0.8+0+0.6+0.5+0.6) = 1.267. If A gets sick first, according to formula (6), the changes of the infection probability of A, B and C with time are shown in table2.

**Table1:** Interaction frequency matrix among three individuals

| Interaction_matrix | A | B | C |
| --- | --- | --- | --- |
| A | 0 | 0.8 | 0.5 |
| B | 0.8 | 0 | 0.6 |
| C | 0.5 | 0.6 | 0 |

**Table2:**
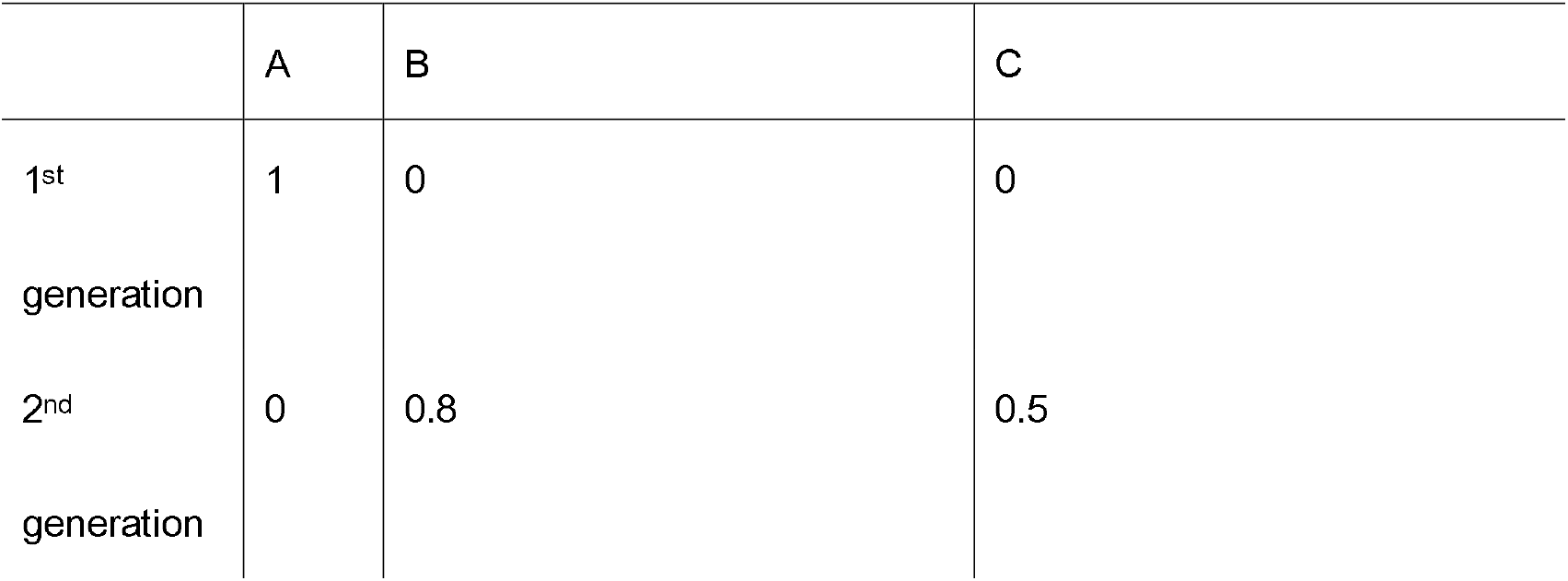
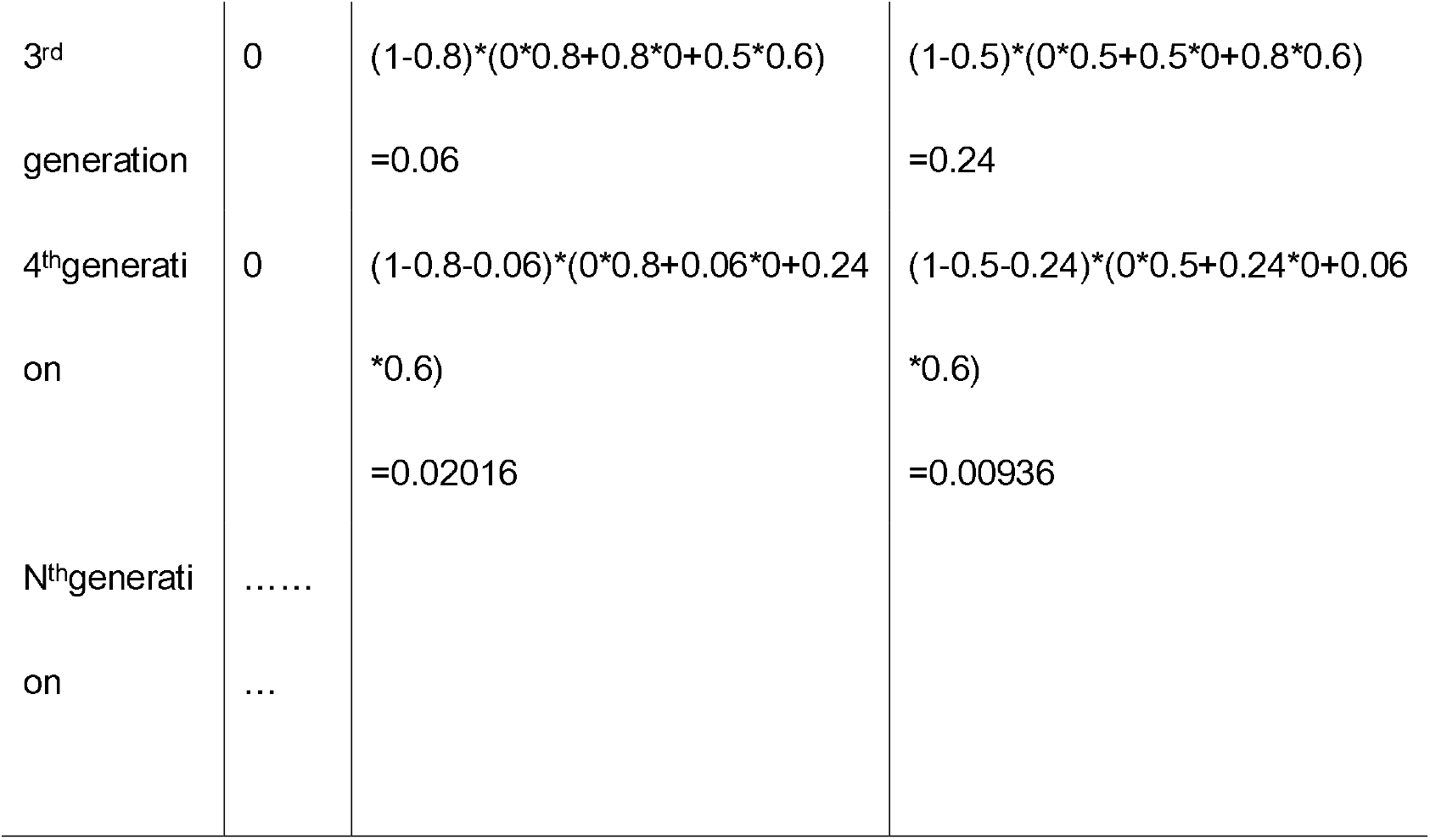
Infection probability of three individuals at different time point

|  | A | B | C |
| --- | --- | --- | --- |
| 1<sup>st</sup> generation | 1 | 0 | 0 |
| 2<sup>nd</sup> generation | 0 | 0.8 | 0.5 |
| 3<sup>rd</sup><br>generation | 0 | $(1-0.8)*(0*0.8+0.8*0+0.5*0.6)$<br>=0.06 | $(1-0.5)*(0*0.5+0.5*0+0.8*0.6)$<br>=0.24 |
| 4<sup>th</sup>generati<br>on | 0 | $(1-0.8-0.06)*(0*0.8+0.06*0+0.24$<br>*0.6)<br>=0.02016 | $(1-0.5-0.24)*(0*0.5+0.24*0+0.06$<br>*0.6)<br>=0.00936 |
| N<sup>th</sup>generati<br>on | .....<br>... |  |  |

Our Bayesian model is a continuous model, that is, the infection of each individual in a specific period of time is treated as a probabilistic problem, rather than a simple infected or uninfected state which would be represented as a Boolean number. The number of infected patients in a population at a certain time point is the sum of the infection probabilities of each individual. When the population size scales up to certain level, this probability could better reflect the actual epidemic dynamic.

### 2.2 The prediction capacity of Bayesian model is significantly better than that of *SIR* model

We expand this model to 10,000 people. We randomly assign the coordinates of these 10,000 people to the square zone with X direction [0-250] and Y direction [0-250]. Using formula (3), we can calculate the contact matrix of the population. When *c1*=0.8, *c2*=5 and *n*=4, according to formula (4), the initial virus reproduction number *R*_*0*_ = 2 can be calculated. We use different models to predict its epidemic curve, and the results are shown in Figure 1.

**Figure1 :**
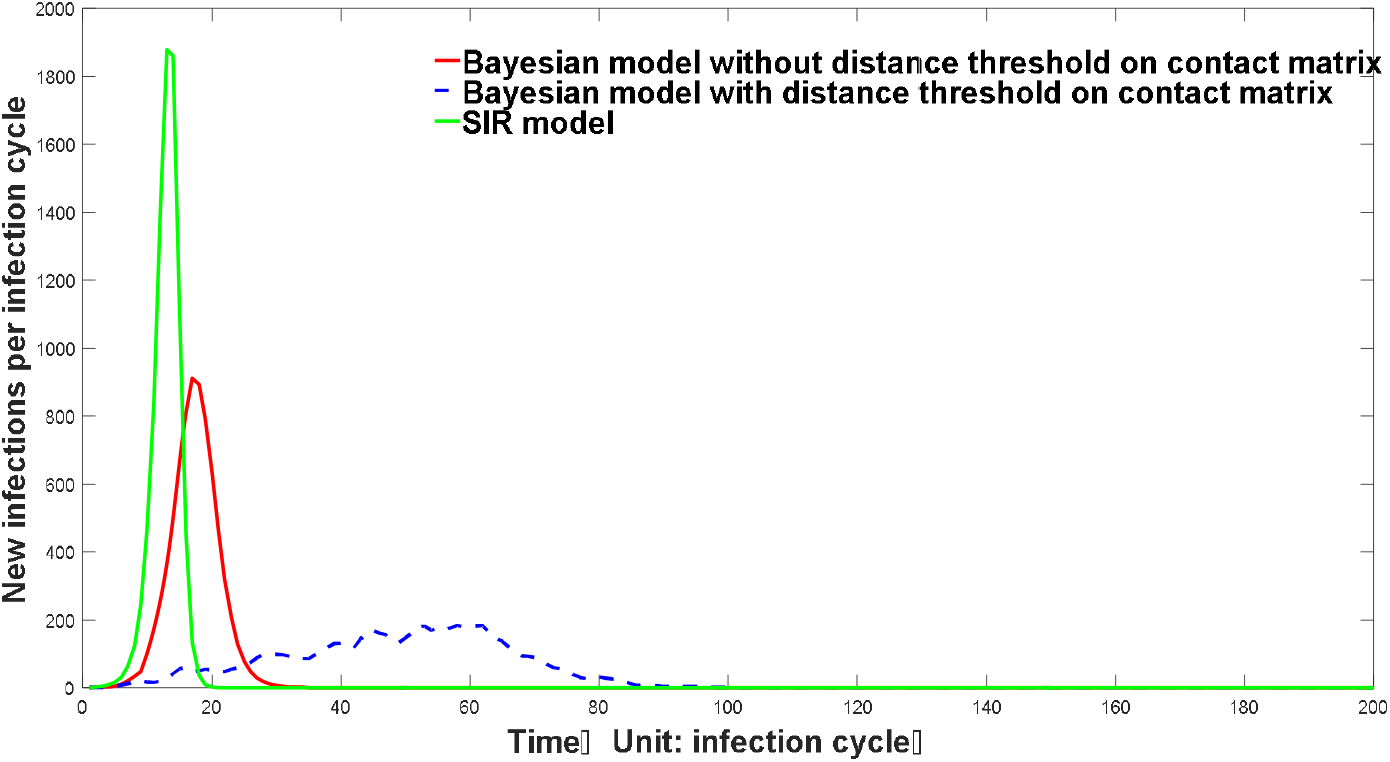
Epidemic trend predicted by three different model

It can be seen from Figure 1 that the early rising trend of the infection curve predicted by *SIR* model is very steep, while the Bayesian model considering population contact is relatively mild. Our Bayesian models are divided into two types, one is the constraint model of population contact distance, that is, contact probability of all people is inversely proportional to the fourth power of the distance between them, that is, the curve represented by the red curve. The other is a model with discontinuous population contact frequency, that is, within a certain distance range, the contact frequency of all people is inversely proportional to the fourth power of the distance between them, and the contact frequency of individuals beyond this distance threshold is 0, which is indicated by blue dotted line. The individual contact frequency of the real population may be between the two. It can be seen from the figure that the epidemic trend curve generated by Bayesian model with population diffusion constraint rises very slowly, and the epidemic growth of Bayesian model without population diffusion constraint is significantly milder than that of *SIR* infection model. The infection curve predicted by Bayesian model is closer to the real infection curve. The results predicted by *SIR* model may deviate greatly from the real situation. One important factor leading to this deviation is that this model does not comprehensively consider the time and space factors of virus infection, especially the influence of population contact matrix brought by population geographic distribution on the overall infection curve. To give a simple example, traditional infectious disease models often predict a very steep epidemic rising trend. On the other hand, using SIR model to force curve fitting often results in a very small number of susceptible people, which is unrealistic compared with the actual situation. The main reason for this phenomenon is that the traditional ordinary differential equation model presumes that infected people have infinite flow and diffusion ability. Thereafter in a very large population, when only a small number of people are infected, the change of *R*_*0*_ can almost be ignored, so the early epidemic prediction is often exponential. However, the actual situation is not the case. Even without any means of prevention and control, the growth of the epidemic will never be exponential, which is mainly due to the spatial effect of diffusion. When a person is infected with a virus, he will give priority to causing infection to nearby people instead of equally causing infection to people in random areas. This mode of transmission will lead to a significant decrease in *R*_0_ value even when only a small number of people are infected. The spatial effect of this epidemic spread can be well reflected by our Bayesian model. As we will see later, the infection curve predicted by Bayesian model can reflect the complex and real epidemic fluctuation when multiple features such as population spatial distributions are considered. At the same time, our model can predict and track the hot spots of epidemic situation, as shown in video 1, and can effectively simulate the dynamic process of epidemic situation at different geographic and time scales.

### 2.3 There is no simple derivation relationship between the virus reproduction number *R*_*0*_ and the final herd immunity threshold, and it may require a higher population infection ratio to achieve the complete extinction of the epidemic through natural herd immunization

As can be seen from Figure 2, based on different methods, the threshold of group immunity predicted by different *R*_*0*_ value is significantly different. The threshold of group immunity predicted by SIR model is the highest, while the value of group immunity predicted by our Bayesian model is significantly higher than that predicted by *R*_*0*_ directly. For Covid-19, assuming its *R*_*0*_equal to 3, the threshold of herd immunity predicted by simple Bayesian model is above 95%, which is significantly different from the 66.6%(1-1/ *R*_*0*_) presumed by using *R*_*0*_ value. The correct prediction of group immunity threshold plays an important role in guiding the formulation of public policies such as vaccination. The herd immunity threshold predicted based on the simple Bayesian model ignores many factors, such as virus variation and individual immunity differences, so the predicted group immunity threshold is not necessarily accurate. However, a simple Bayesian model can reflect a problem, that is, the simple method of inferring group immunity threshold based on *R*_*0*_ value is inaccurate and unreliable. Although there is a significant positive correlation between virus reproduction coefficient *R*_*0*_ and group immunity threshold, the presumed relationship which represents as *Threshold = 1-1/ R*_*0*_ is not valid. The threshold of herd immunity deduced by this assumption is often too low.

**Figure2 :**
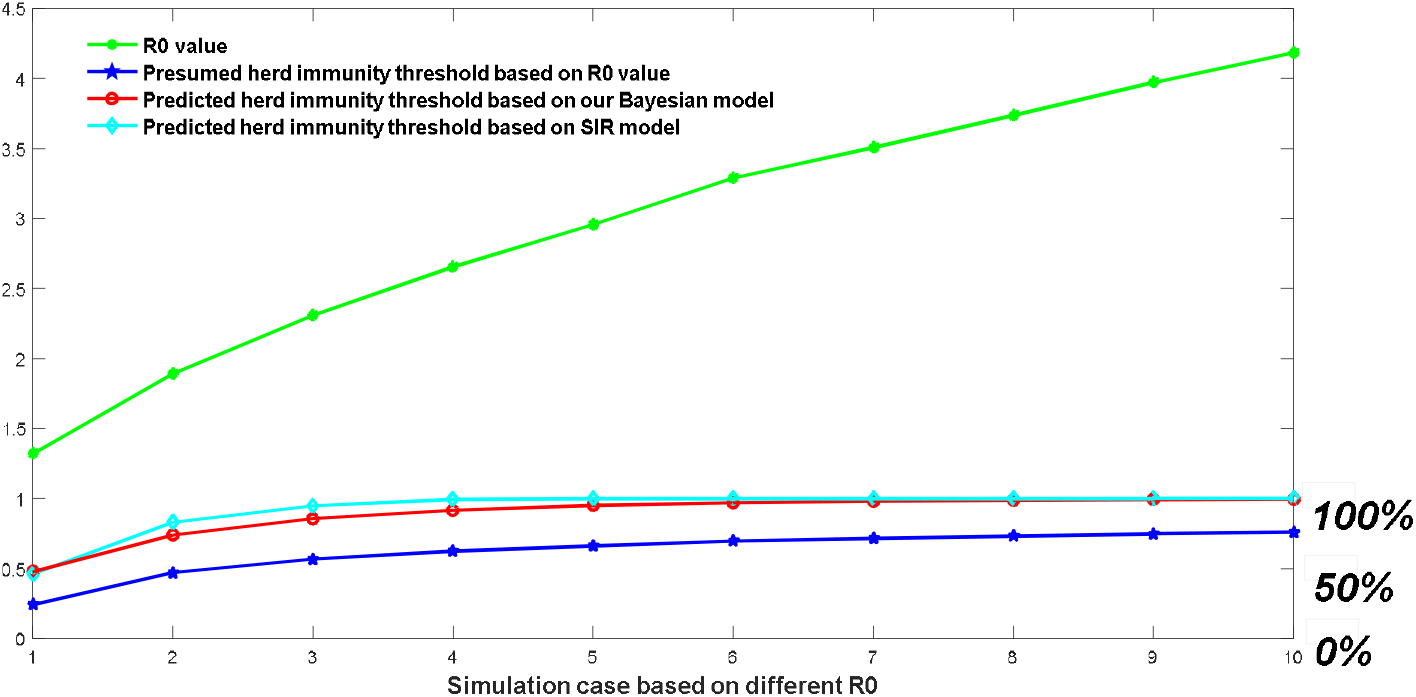
Herd immunity threshold predicted by three different approaches

We further studied the influence of vaccination rate on the final number of infections. We assumed that the vaccination was completed instantaneously, and the effect of vaccination on our model was equivalent to indirectly reducing population density. For example, assuming that the vaccine is 100% effective, a 70% vaccination rate is equivalent to 3,000 people randomly distributed in the original area instead of 10,000 people. The relationship between the vaccination rate and the final number of infected people is shown in figure 3.

**Figure3 :**
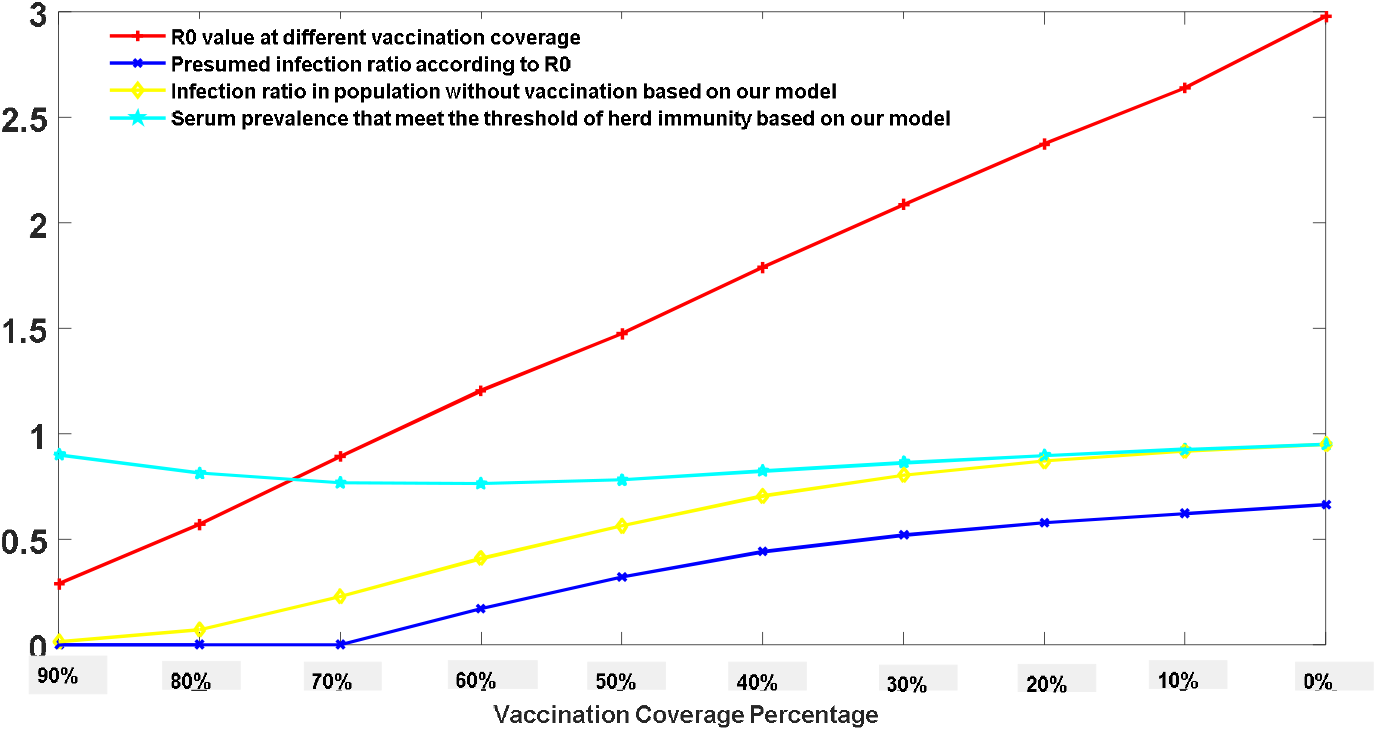
Predicted infection probability at different vaccination coverage using a simple Bayesian model

It can be seen from fig. 3 that for a virus with reproduction number*R*_*0*_ = 3, on the premise of 100% vaccine effectiveness, there is a linear negative correlation between vaccination rate and reproduction number*R*_*0*_ after vaccination. Yellow curve indicates the infection ratio of uninoculated people predicted by Bayesian model. The results indicate that 90% vaccination rate will cause 1.2% infection probability of the remaining 10% population; 80% vaccination rate will cause 7.2% infection probability in the remaining 20%; 70% will cause 22.8% infection probability in the remaining 30% and 60% will cause 40.9% infection probability in the remaining 40%. Since there is no available method in guiding the public to decide what vaccination coverage percentage is the best, the Bayesian model provides a mathematical but reasonable way in this optimization problem.

### 2.4 A more realistic situation: a Bayesian model considering virus mutation, natural attenuation of antibodies, population geographic distribution, population age structure, asymptomatic infections and many other features

For a realistic infection situation, we often need to consider more variables, such as the influence of virus mutation and antibody attenuation effects, the effects of population regional distribution, and the impact of population age structures on the epidemic development. At the same time, a very important aspect is that we must consider the dose effect on infection probability, that is, the relationship between the infectivity of patients and the initial amount of invading virus. A notable phenomenon in COVID-19 infection is the emergence of a large number of asymptomatic patients. Moreover, an interesting circumstance is that the proportion of serum prevalence is much higher than the reported number of infected people ^[9]^. Experiments have confirmed that the severity of patients’ symptoms is positively correlated with viral load in vivo ^[10,11]^. Our model holds that different infections will possess different transmission potentials. The definition of ‘infection’ in our model is based on the existence of antibodies. For the same individual, when small amount of virus invades, its infectivity caused by infection will be reduced. A correction function (equation (10)) added to transform its infection possibility into the transmission potential after infected. We simulated the epidemic dynamics of a specific region with population distributed at four different cities. Most people in different cities have no direct contact opportunity, except a few of them. These few people become the links that connect the interactions in different regions. We assume that the mutation constant of virus is 0.002, the attenuation constant of antibody is 0.005, the relationship between infection occurrence and age follows equation (9), and the correction relationship between infectivity and contact probability follows equation (10). The specific parameters are provided in the supplementary materials, the results are shown in figure4.

Video 1: The forecasted epidemic geographic distribution using a multi-factors Bayesian model

From Figure 4.A, we can see that although the public prevention polices could significantly affect the development of the epidemic situation, the population geographic distribution is also an important factor, or even a dominant factor in driving the trend of epidemic. Under a relatively stable public prevention strength, the spatial distribution of population will lead to the wave-like epidemic fluctuations and display multiple peak points. This trend has been fully reflected in countries and regions suffering COVID-19 epidemic. Therefore, when we forecast epidemic development, we need to consider the spatial and geographical factors. The short-term decline does not necessarily represent the overall decline of the epidemic situation, but may be the signal of epidemic spreading from one region to another. From figure 4B to figure 4I, we can see the spatial migration of infection hotspots more clearly. For example, at time point B, the infected people mainly concentrated in city 1, while at time point C, the infection hotspot moved to city 2, and the epidemic situation in city 1 subsided to a certain extent. Another important function of Figure 4.A is to evaluate the impact of virus mutation and antibody decay on the epidemic development. The function of virus mutation and antibody decay we used is a simplified function, which lacks sufficient data support, but it can roughly mimic the actual situation to a certain extent. From our modeling results, we must be prepared to coexist with Covid-19 for a long time, because there is a possibility, at least in theoretical level that the virus may not be completely eliminated by natural immunization or vaccination. Because of COVID-19’s natural attributes, high mutation rate and the existence of antibody fading effect which means antibodies produced by human body are fading away through time, the future epidemic distribution may not have hotspots, but will be randomly distributed around the world with a relatively small probability, and Covid-19 will become a wild-distributed virus. Specifically, as shown in fig. 4G, at time point G when a complete herd immunization cycle has been realized, the epidemic may be re-occured for the second time in city 1. At that time, the epidemic situation is characterized by the lack of concentrated hotspots, and a wild-distribution with small probability. As our whole human society, the complexity of its population and spatial distribution far exceeds the scale of our model which would provide more favorable breeding grounds for the evolution of viruses, so the probability of reinfection will greatly increase. The epidemic recurrence was already on its way even after high vaccination coverage. This has already been confirmed by the third epidemic wave in Britain starting from June,2021.

**Figure4.A :**
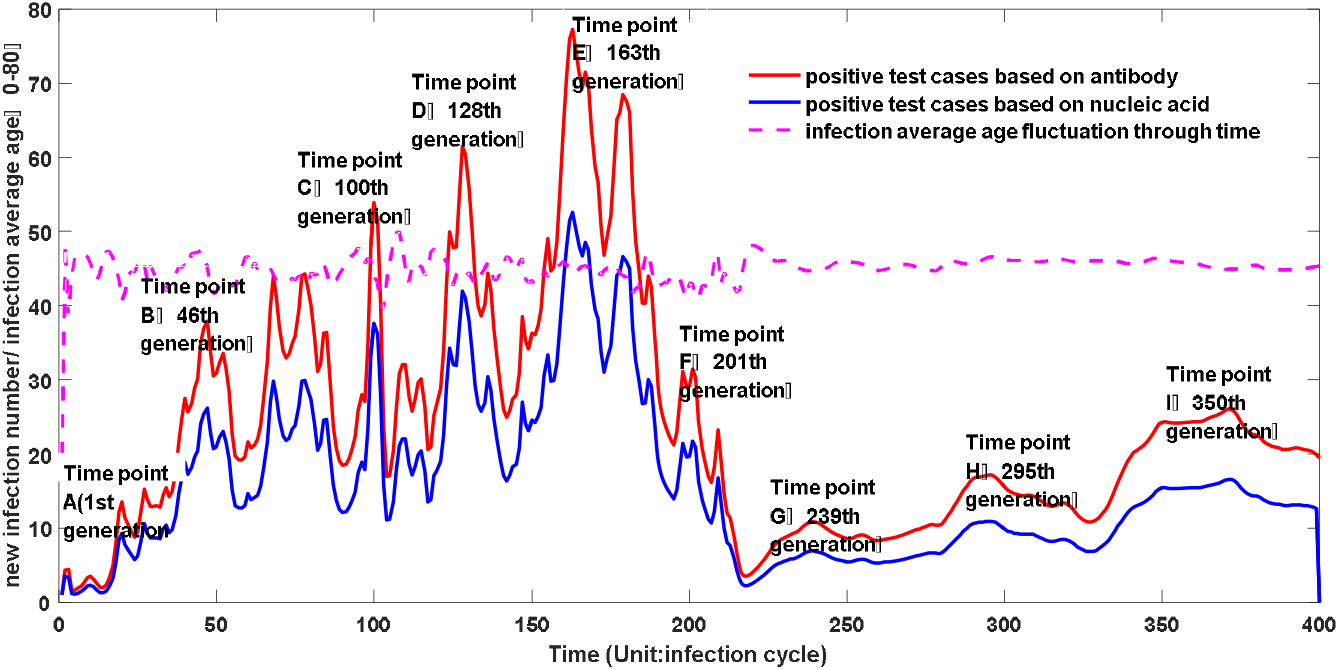
Epidemic trend predicted by Bayesian model with the consideration of multiple factors

**Figure4.B :**
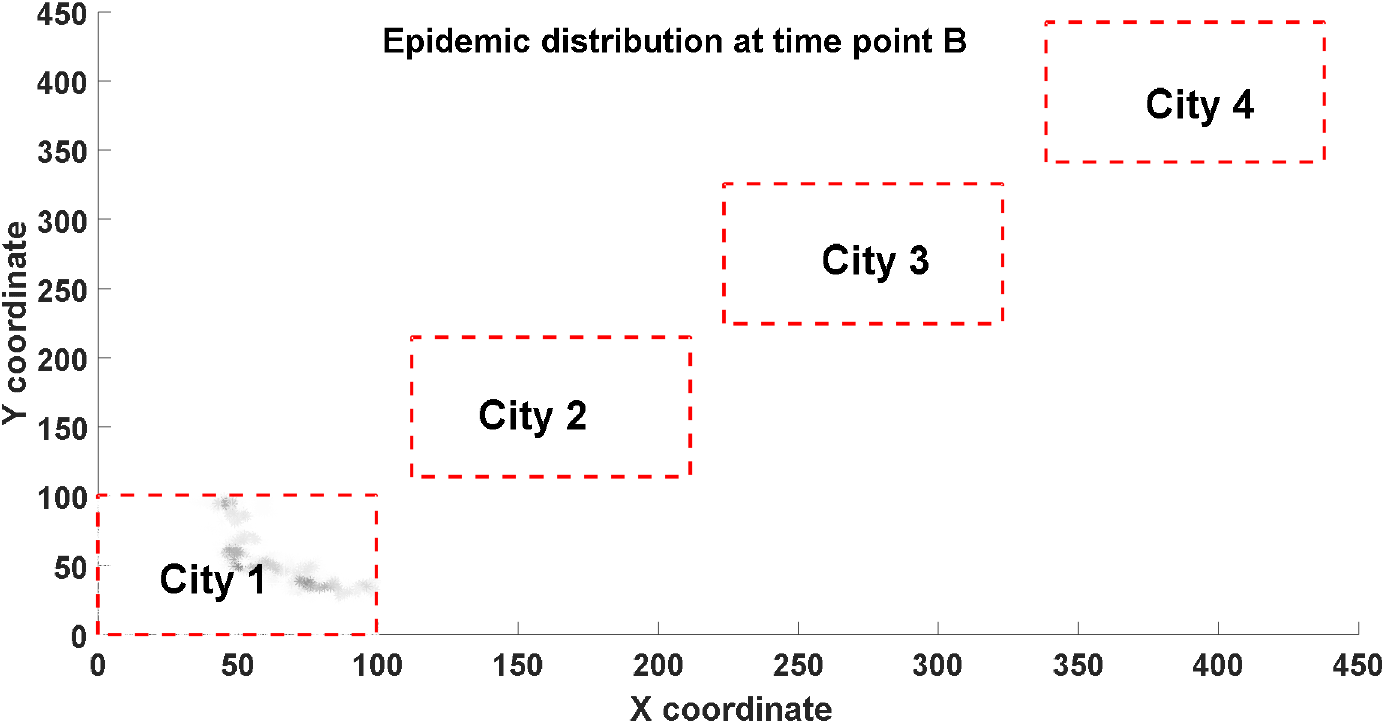
The predicted epidemic distribution at time point B using a multi-factors Bayesian model

**Figure4.C :**
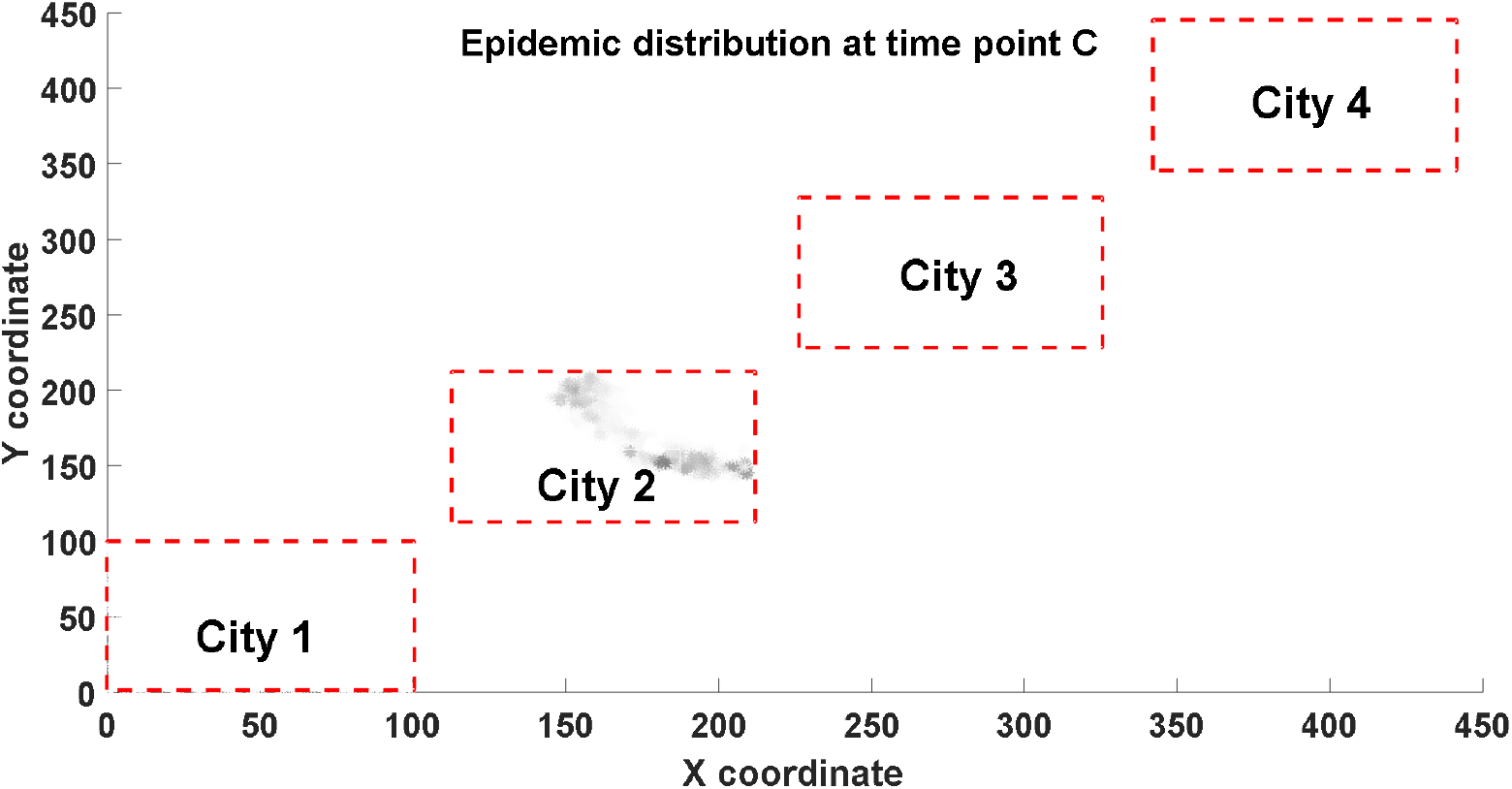
The predicted epidemic distribution at time point C using a multi-factors Bayesian model

**Figure4.D :**
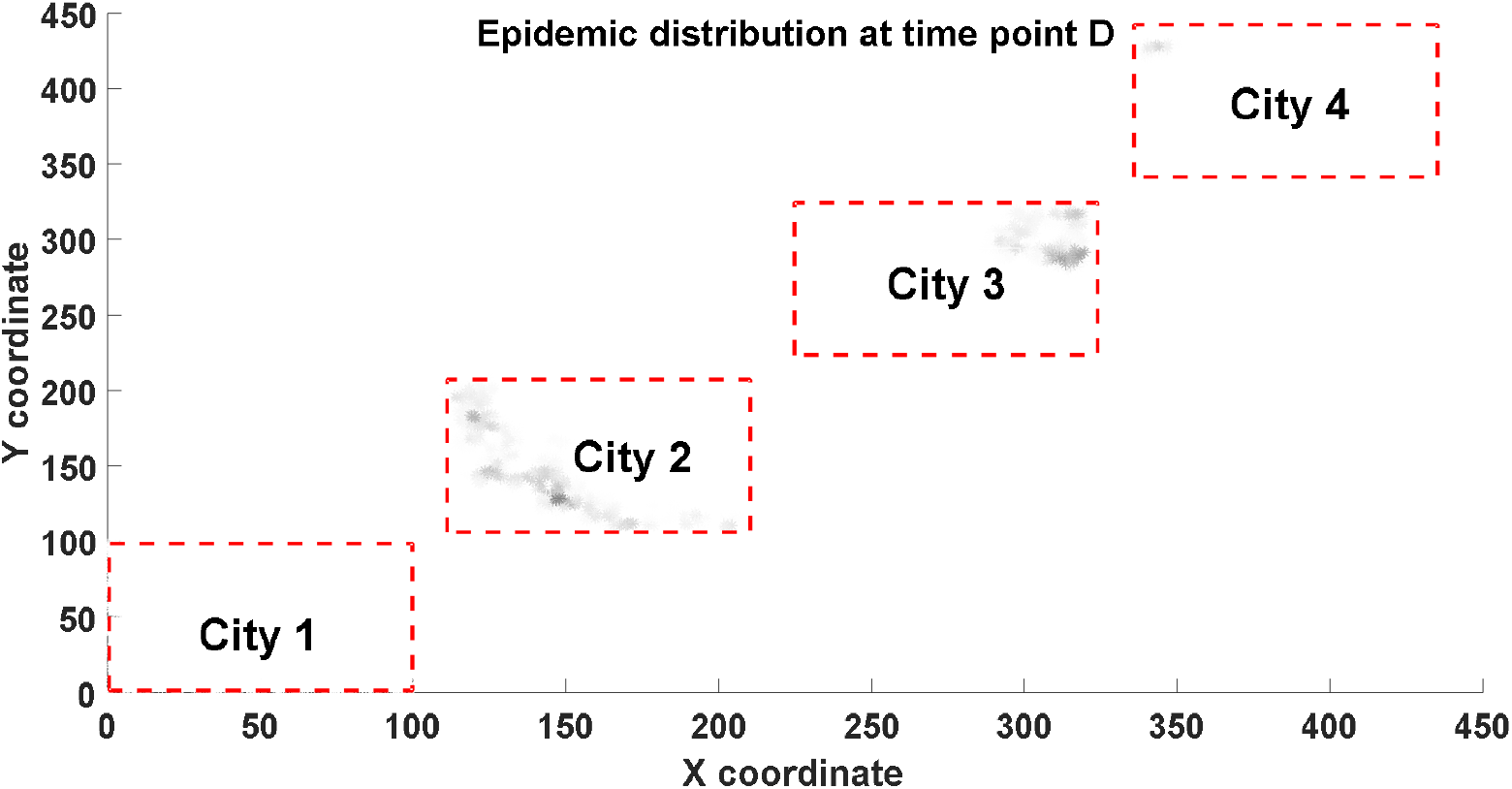
The predicted epidemic distribution at time point D using a multi-factors Bayesian model

**Figure4.E :**
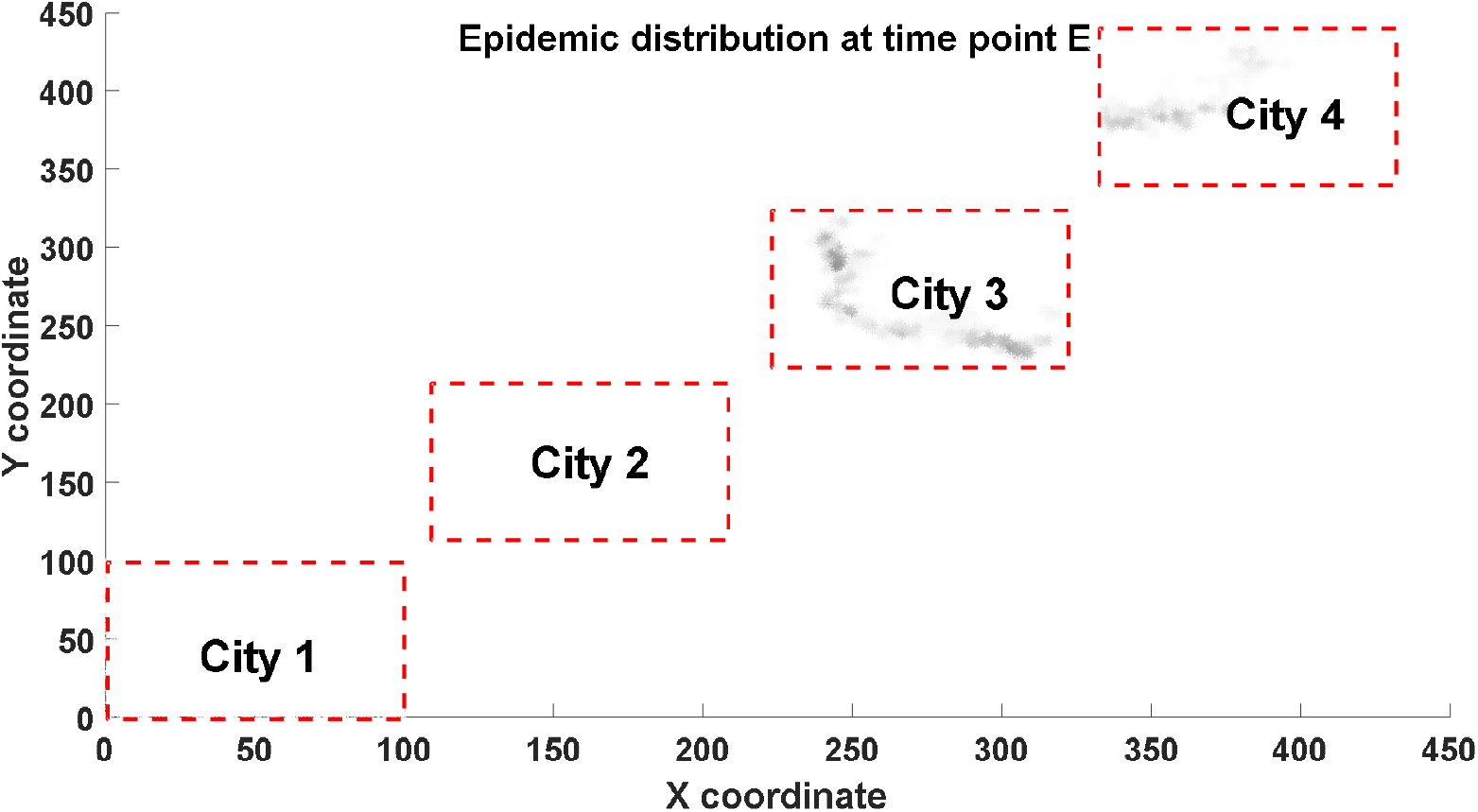
The predicted epidemic distribution at time point E using a multi-factors Bayesian model

**Figure4.F :**
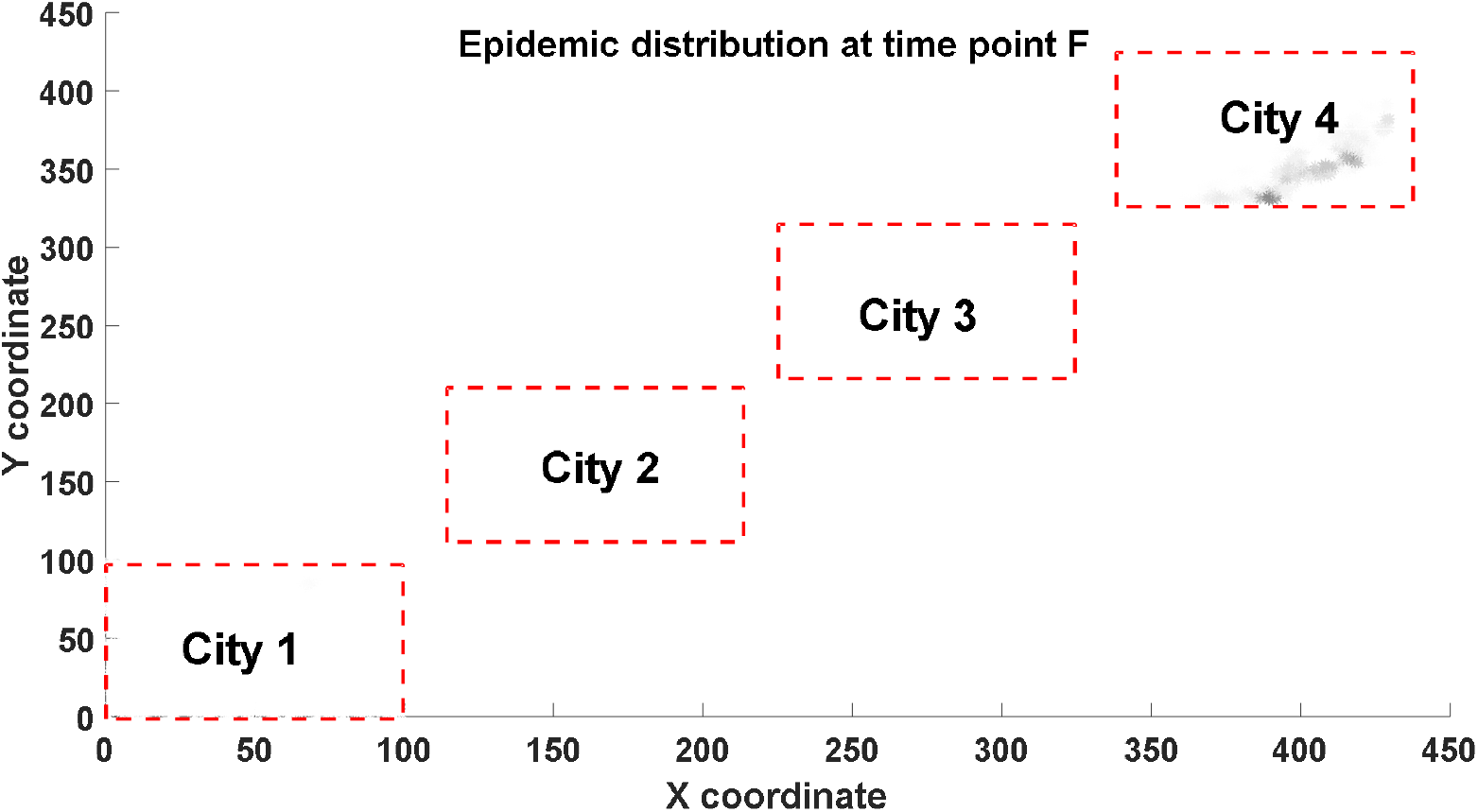
The predicted epidemic distribution at time point F using a multi-factors Bayesian model

**Figure4.G :**
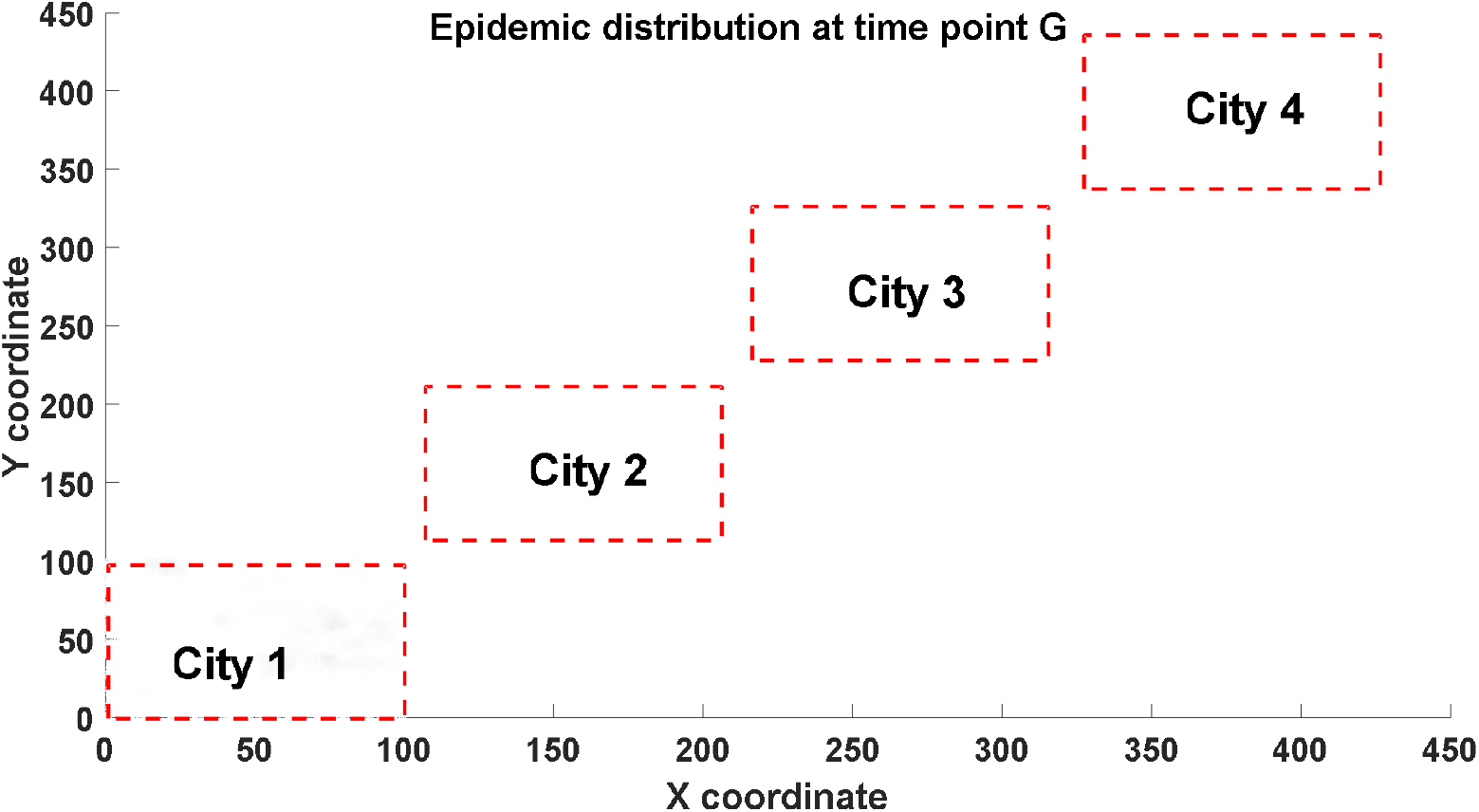
The predicted epidemic distribution at time point G using a multi-factors Bayesian model

**Figure4.H :**
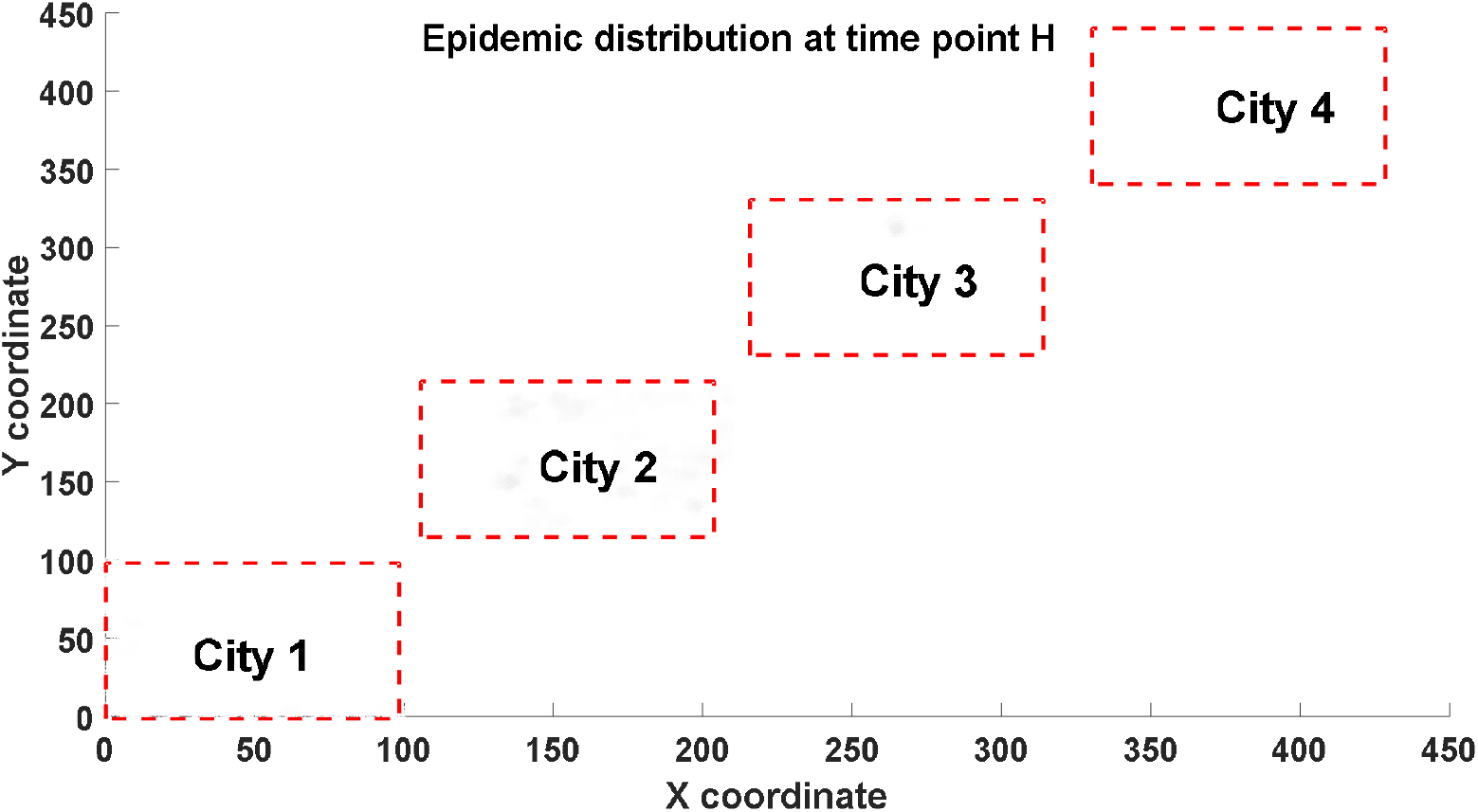
The predicted epidemic distribution at time point H using a multi-factors Bayesian model

**Figure4.I :**
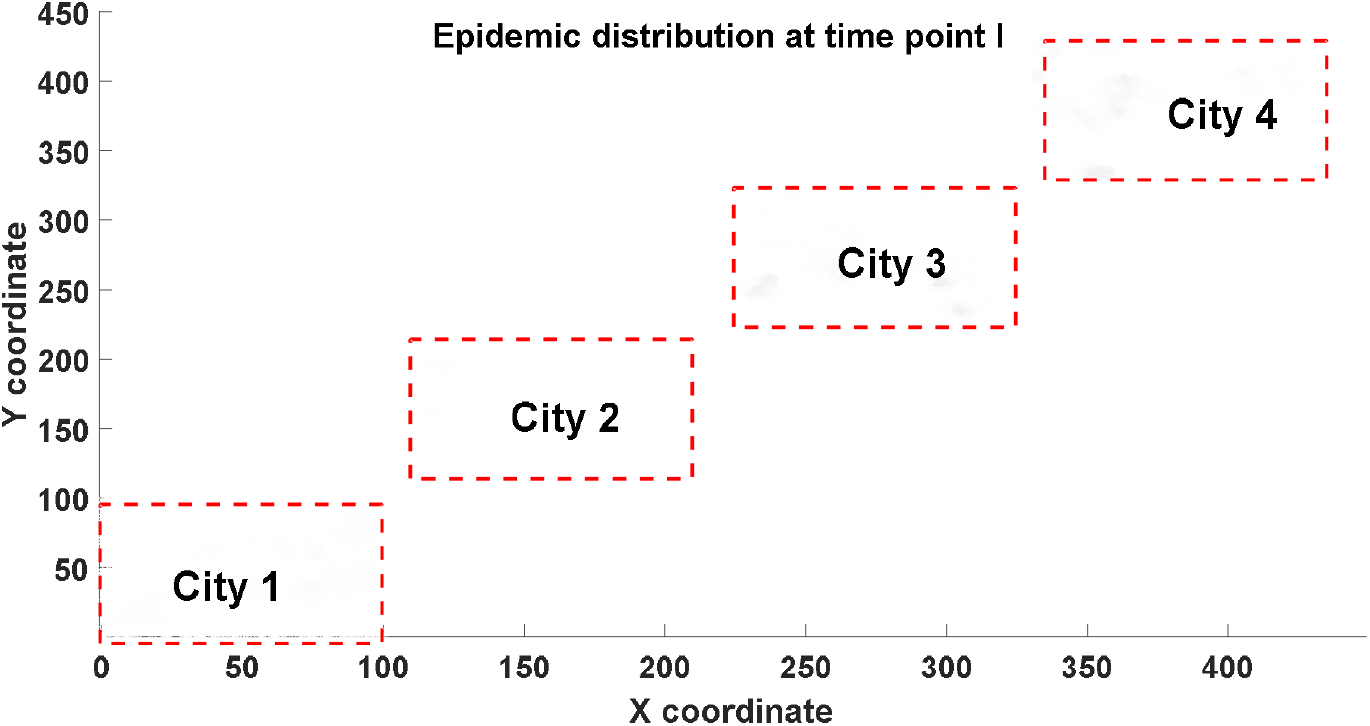
The predicted epidemic distribution at time point I using a multi-factors Bayesian model

At the same time, we can predict the threshold of group immunity in the real situation through parameter estimation. This threshold does not have simple correlation with *R*_*0*_and is closely related to population age structure and population contact matrix. For simulation described above, the calculated virus reproduction coefficient *R*_*0*_ is 2.1175, which corresponds to the traditional group immunity threshold of 52.7%. However, the actual serum prevalence reached 66.2% after a natural herd immunization cycle (1-210 generations), among which 1.3% of infected people had a second infection. Similar phenomenon has been reported by a serum prevalence study of Iran, indicating at epidemic hotpots, the antibody positive rate has further exceeded the herd immunity threshold derived directly from *R*_*0*_. Specifically, Nazemipour and colleagues stated that 72·6% seroprevalence in Rasht city which did not follow the presumed threshold of herd immunity ^[17]^.

Using the Bayesian model, we can estimate the proportion of the population needed to complete the first round of group immunization, and at the same time, we can calculate the serum prevalence at different age group. We also noticed another interesting phenomenon. As shown by the purple dotted line in Figure 4, the average age of infection does not engage a significant alternation, which means that the infectivity of virus to different age groups will not change in the spatial diffusion process. The change of the average infection age during the epidemic may be caused by other factors, such as the change of exposure frequency caused by age factors or some intrinsic features at virus genome level.

## Discussion

John von Neumann once famously said”With four parameters I can fit an elephant, and with five I can make him wiggle his trunk.” By this he meant that one should not be impressed when a complex model fits a data set well. With enough parameters, you can fit any data set. Compared with the traditional *SIR* method, the *SIR* method with increased parameters, including various *SIRD* (Susceptible, Infected, Recovered, Death) model, *SEIRD* (Susceptible, Exposed, Infected, Recovered, Death) model and so on, all can achieve good fitting results. Nevertheless, those models all fall into the trap of pure mathematical fitting. Using multi-parameters, one can produce better fitting results. But multiple parameters can also bring several critical problems: First, the solution of parameters is not unique. Second, due to the lack of strong physical mechanism, it does not have a good prediction effect, which has been confirmed in many early studies of COVID-19 epidemic ^[12-16]^. None of these models can accurately predict the inflection turning point of the epidemic, let alone the repeated fluctuations of the epidemic. Therefore, we should try to abandon the traditional idea of using parameter fitting to predict the epidemic situation, and develop a method that can integrate more specific, more realistic and more complex information into the model, which could bring more reliable and accurate prediction results.

Based on this idea, we established a Bayesian model of virus infection for the first time. Our model can effectively consider the impact of the actual contact probability of population on the epidemic development. The population contact probability matrix can be roughly calculated according to the population spatial distribution. Besides that, we can also integrate in-vivo individual contact frequencies reflected in real situation into our model. Compared with *SIR* and other ordinary differential equation system models, Bayesian model can integrate more information, such as the contact frequency of different individuals, which is closely related to spatial location and individual relationship. Meanwhile, it can comprehensively consider the virus mutation effect, antibody attenuation effect, population age structure, etc. All of these advantages enable this model a more powerful prediction capacity, which can not only predict the epidemic dynamics through time, but also detect the epidemic hotspots distribution at different time. If we are able to access an authentic data for analysis, we can fit the model parameters of different regions more accurately. Using these parameters, we can effectively forecast the spatial and temporal trend of epidemic situation and predict the threshold of herd immunity. What we have to reiterate is that the herd immunity threshold does not have a simple relationship with its *R*_*0*_. The actual herd immunity threshold might be significantly higher than the presumed one derived from*R*_*0*_. This finding might have a significant influence on the future public decision which indicates a higher vaccination coverage need to be reached in order to meet the threshold of herd immunity.

There are still many defects in our model. For example, the computational cost is proportional to the square of population size. Although our model has a great potential to stimulate with more realistic statistical data, we have not applied it to an in-vivo population contact situation. The future research mainly includes improving the algorithm efficiency, integrating in-vivo data to obtain more reliable parameters, and verifying the reliability of this method in the analysis of real cases.

## Data Availability

All data and codes in this reprint are available through the link below.

https://github.com/zhaobinxu23/A-Continuous-Bayesian-Model-for-the-Simulation-of-SARS-CoV-2-Epidemic

## Supplementary materials

Matlab codes can be accessed through the following link: https://github.com/zhaobinxu23/A-Continuous-Bayesian-Model-for-the-Simulation-of-SARS-CoV-2-Epidemic

